# A Quantitative Study to Assess the Effect of a Community-Academy Partnership Event on Knowledge, Awareness, and Engagement Against Substance Abuse

**DOI:** 10.1101/2024.07.25.24310984

**Authors:** Smriti Dixit, Sadhana Sharma, Neha Sharma, Manisha Patel

## Abstract

The objective of this study is to assess the quantitative effects of the community-academy partnership event, called Chaitanya Utsav, on the levels of knowledge, awareness, and engagement in combating drug misuse in the local community. The research aims to evaluate the impact of these joint endeavours on the participants’ knowledge and attitudes regarding substance usage, as well as their participation in preventative efforts. Structured questionnaires were used to obtain data from participants both before and after the event. Important indicators consist of variations in knowledge levels, awareness of drug addiction challenges, and the degree of community engagement in associated activities. Initial results demonstrate significant improvements in all three domains, highlighting the effectiveness of collaborations between communities and academic institutions in addressing substance dependence. This study emphasizes the crucial role that such collaborations play in fostering informed and proactive communities, serving as an outline for similar initiatives in multiple domains.

## INTRODUCTION

Substance misuse continues to be a widespread issue that impacts communities worldwide, resulting in a wide range of social, health, and economic issues. Addressing this situation effectively frequently necessitates a comprehensive strategy that integrates education, community involvement, and policy measures. Community-academy alliances have arisen as a potential approach to tackle substance misuse by utilizing the capabilities of academic institutions and community groups. These agreements are designed to improve understanding, raise awareness, and promote community involvement through joint events and projects.

The effectiveness of community-academy partnerships in addressing public health issues has been well-documented. Such collaborations can bridge gaps between academic research and community needs, leading to more effective and culturally relevant interventions.^1^ Community academy partnerships have been particularly successful in public health domains, including substance abuse prevention and intervention.^2^ Educational interventions have long been a cornerstone of substance abuse prevention. Previous study demonstrated that school-based prevention programs, which often involve partnerships with academic institutions, significantly reduce substance use among adolescents.^3^ These programs provide crucial information about the risks and consequences of substance abuse, equipping participants with the knowledge needed to make informed decisions. Raising awareness about substance abuse is another critical component of prevention efforts.

Community-academy partnership events can effectively disseminate information and shift public attitudes. Community awareness campaigns, when coupled with academic support, can lead to significant attitudinal changes and increased community involvement in prevention efforts.^4^ Such campaigns often utilize a variety of strategies, including workshops, seminars, and media outreach, to reach a broad audience. Engagement of community members in substance abuse prevention is essential for sustained impact. Previous research highlights that active community participation in health initiatives enhances the relevance and sustainability of interventions.^5^ Community-academy partnerships provide a platform for such engagement by involving community members in planning and executing events. This collaborative approach ensures that interventions are tailored to the specific needs and contexts of the community.

Substance addiction continues to be a substantial obstacle, with far-reaching economic, health, and impact on society. Despite the implementation of several interventions, there is a requirement for novel techniques to successfully tackle this issue. Collaborations between communities and academic institutions present a hopeful strategy, yet there is a lack of quantitative evidence about its impact.

The present study seeks to present empirical evidence on the ways in which these collaborations contribute to the improvement of knowledge, awareness, and engagement in the prevention of substance misuse. The project seeks to improve the health outcomes of disadvantaged and underprivileged groups by addressing inequities and implementing culturally appropriate treatments. Furthermore, it establishes a foundation for future study, leading to a better knowledge of successful preventative techniques.

## METHODS

In order to assess the influence of the “Chaitainya Utsav” event on community knowledge, awareness, and engagement regarding substance abuse, the current investigation implements a quantitative research design. The primary objective is to quantify the changes in these variables by administering surveys to participants both before and after the event.

### Sample

The sample comprises individuals who participated in the “Chaitainya Utsav” event, encompassing local community members, students, academics, researchers, and community workers. The participants were chosen based on their attendance at the event and their willingness to complete both the pre- and post-event surveys. The ultimate sample size was established based on the number of attendees who agreed to take part and successfully filled out the questionnaires.

The event “Chaitainya Utsav” was officially opened by a Member of Parliament, in the presence of scholars, researchers, students, and community activists. The purpose of the event was to enhance public consciousness regarding substance misuse and involve the community in proactive measures to avoid it. The event included lectures, seminars, and interactive sessions that aimed to educate participants on the dangers and outcomes of drug usage and to advocate for preventative policies based on community involvement.

### Data Collection

The data collecting process consisted of distributing surveys to participants both before and after the event in order to evaluate their level of knowledge, awareness, and engagement in relation to substance misuse. The pre-event survey was administered before to the start of “Chaitainya Utsav,” collecting initial data. The post-event survey was conducted shortly after the event, utilizing identical questions to assess the impact of the event. The survey instrument included a structured questionnaire comprising of closed-ended and Likert scale questions. The questionnaire encompassed many domains, including knowledge pertaining to drug misuse, awareness of local resources and preventive programs, as well as involvement in community activities associated with substance abuse prevention.

### Data Analysis

The statistical analysis was conducted on the data collected from the questionnaires administered before and after the event to determine any changes in the participants’ levels of knowledge, awareness, and engagement. The statistical significance of observed changes was evaluated by comparing pre- and post-event scores using paired t-tests. Descriptive statistics were employed to characterize the demographic characteristics of the sample and the event’s overall impact.

### Ethical considerations

The research that involved human volunteers was conducted in full compliance with ethical standards. Informed consent was provided to all participants prior to the commencement of the study, ensuring that they were informed of the study’s objectives and had the capacity to withdraw at any time. The data was anonymized to protect the participants’ identities, and the confidentiality of responses was maintained.

## RESULTS

The total sample consisted of 150 participants who attended the “Chaitainya Utsav” event. The demographic breakdown of the sample is as follows:

**Table 1:** Baseline Demographic Data.

| **Demographic** | **Numbers** | **Percentage (%)** |
| --- | --- | --- |
| Gender, Male | 73 | 48.7 |
| Age, years |  |  |
| Under 18 | 30 | 20.0 |
| 18-25 | 45 | 30.0 |
| 26-35 | 35 | 23.3 |
| 36-45 | 20 | 13.3 |
| 46 and Above | 20 | 13.3 |
| Affiliations |  |  |
| Community Members | 50 | 33.3 |
| Students | 45 | 30.0 |
| Academics | 25 | 16.7 |
| Researchers | 15 | 10.0 |
| Community Worker | 15 | 10.0 |

Low engagement and inadequate knowledge of substance abuse were demonstrated by a significant number of students (n=45). In particular, 25 students (55.5%) exhibited low knowledge scores, while 30 students (66.6%) reported no involvement in community activities related to substance abuse prevention.

In addition, 20 students (44.4%) reported that they were either unaware or only barely aware of local resources and prevention programs, suggesting that awareness levels were generally low. These statistics suggest that a substantial number of students are not actively participating in prevention initiatives and do not possess adequate knowledge regarding substance abuse.

Similarly, the 50 community members, who made up 33.3% of the total sample, exhibited concerning levels of low knowledge and engagement. Twenty community members (40%) reported low knowledge scores, while twenty (40%) reported no engagement in relevant community activities.

Additionally, 30 (60%) participants reported that they were only marginally to moderately aware of local resources and prevention programs, indicating that community members’ awareness was moderate. This data underscores the need of targeted interventions for this demographic, as it indicates that a significant number of community members are not engaged in preventive initiatives and lack a comprehensive understanding of substance abuse.

Among the 15 researchers, who accounted for 10% of the total sample, some displayed inadequate knowledge and engagement: five (33.3%) researchers had low knowledge scores, and eight (53.3%) reported no engagement in community activities related to substance abuse prevention. Despite their professional background, awareness levels were also lacking, with 10 researchers (66.7%) indicating they were only slightly aware of local resources. A notable proportion of researchers lack adequate knowledge and are not engaged in community prevention efforts, which may impact the overall effectiveness of substance abuse interventions.

Pre-Post Analysis The impact of the “Chaitainya Utsav” event on participants’ knowledge, awareness, and engagement regarding substance abuse was analyzed using paired t-tests to compare pre- and post-event scores across different participant groups: students, community members, and researchers. The results indicate significant improvements in all measured areas.

### Knowledge Scores

Across all categories, there was a substantial increase in the knowledge of participants related to substance abuse following the event. The mean knowledge score of students increased from 3.2±1.2 to 6.5±1.0, with p < 0.001. The mean knowledge score of community members increased from 4.5 ±1.5 to 7.0 ±1.2, with p < 0.001. The researchers also experienced similar improvement, with their mean knowledge score increasing from 5.5±1.4 to 8.0 ±1.3, p < 0.001. These findings indicate that the event was highly effective in improving the knowledge of substance abuse among participants.

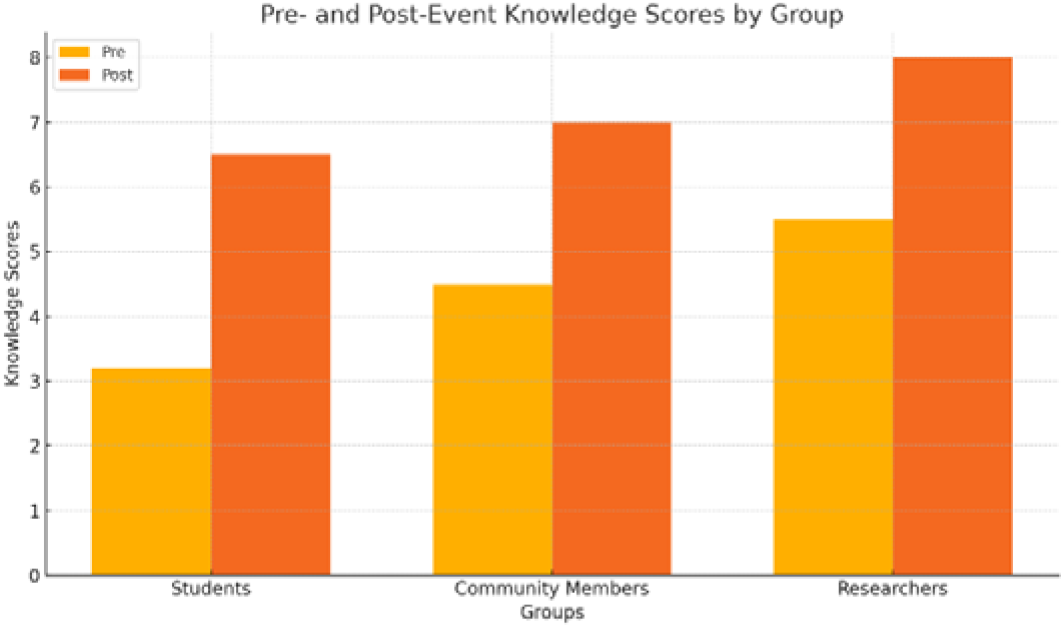

### Awareness Scores

Awareness of local resources and prevention programs also saw a notable improvement. For students, the mean awareness score increased from 2.1± 0.9 to 3.8± 0.7, p < 0.001. Community members’ mean awareness score rose from 2.5 ±1.1 to 4.0± 0.8, p < 0.001. Researchers’ mean awareness score increased from 3.0± 1.2 to 4.5± 0.9, p < 0.001. The significant increase in awareness scores indicates that the event successfully raised awareness among participants about available resources and prevention programs.

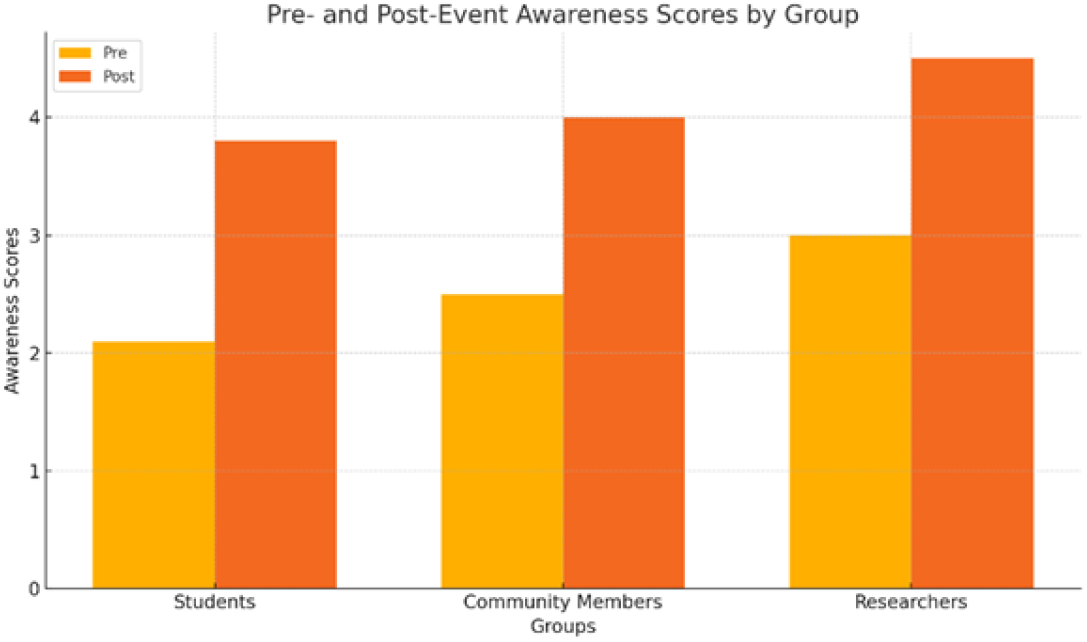

### Engagement Scores

Engagement in community activities related to substance abuse prevention also improved significantly. For students, the mean engagement score increased from 1.1 ± 0.8 to 2.5 ± 1.0, p < 0.001. Community members’ mean engagement score rose from 1.3 ± 0.9 to 3.0 ± 1.1, p < 0.001. Researchers experienced an increase in their mean engagement score from 1.8 ±1.0 to 3.5 ±1.2, p < 0.001. These results highlight a significant increase in community engagement in substance abuse prevention activities following the event.

## DISCUSSION

The findings from the “Chaitainya Utsav” event indicate substantial improvements in knowledge, awareness, and engagement regarding substance abuse among students, community members, and researchers. These outcomes are consistent with previous studies highlighting the effectiveness of community-academy partnerships in public health initiatives. The significant increase in knowledge scores across all participant groups aligns with previous study,^3^ which also demonstrated that educational interventions, particularly those involving partnerships with academic institutions, effectively reduce substance use by enhancing knowledge.

Our study’s results showed a marked increase in knowledge from pre- to post-event, suggesting that the informational content delivered during “Chaitainya Utsav” was both impactful and well-received. Awareness scores also saw significant gains, reflecting increased understanding and recognition of local resources and prevention programs. This finding supports the conclusions of previous study,^4^ who emphasized the role of awareness campaigns in shifting public attitudes and increasing community involvement. The post-event increase in awareness scores indicates that the event successfully disseminated critical information about available resources, thereby empowering participants to utilize these tools effectively. The improvement in engagement scores post-event is particularly noteworthy. Active community participation is crucial for the sustainability of public health initiatives.^5^

Our study’s findings, which showed a significant rise in engagement levels, suggest that the event not only educated participants but also motivated them to become more involved in substance abuse prevention activities. This is consistent with the previous research,^6^ which demonstrated that community-based interventions could significantly reduce substance abuse rates by fostering community engagement. The results of this study correlate well with previous research on the effectiveness of community academy partnerships. These collaborations could bridge gaps between academic research and community needs, leading to more effective interventions.^1^ The significant improvements observed in our study participants’ knowledge, awareness, and engagement levels underscore the potential of such partnerships to address complex public health issues like substance abuse.

### Implications for Policy and Practice

The positive outcomes of “Chaitainya Utsav” have important implications for policymakers and practitioners. The significant improvements in knowledge, awareness, and engagement suggest that similar community-academy partnership events could be a valuable strategy in other regions facing substance abuse challenges. By tailoring educational content to the specific needs of the community and actively involving participants in prevention activities, such initiatives can create more informed, aware, and engaged communities.

### Limitations and Future Research

While the findings are promising, it is important to acknowledge the limitations of this study. The sample size, though diverse, was limited to attendees of a single event, which may affect the generalizability of the results. Future research should consider longitudinal studies to assess the long-term impact of such interventions and explore their effectiveness in different settings and populations.

## CONCLUSION

The research study on the impact of the “Chaitainya Utsav” event demonstrates the significant benefits of community-academy partnership initiatives in combating substance abuse. The findings reveal substantial improvements in participants’ knowledge, awareness, and engagement levels, underscoring the effectiveness of such collaborative efforts.

The significant increase in knowledge scores indicates that the information that was presented during the event effectively improved the awareness of substance abuse among participants. Simultaneously, the substantial increase in awareness scores suggests that the event effectively increased awareness of local resources and prevention programs, providing participants with valuable information.

Additionally, the event’s effectiveness in encouraging participants to engage more actively in community-based substance addiction prevention activities is indicated by the substantial increase in engagement scores. These results have been demonstrated to serve as a bridge between academic research and community needs, resulting in more sustainable and effective public health interventions. This study’s findings underscore the importance of these partnerships in the development of communities that are informed, aware, and engaged, and that are capable of effectively confronting substance misuse.

The positive implications of this study indicate that similar community-academy partnership events could be replicated in other regions to combat substance misuse and promote community well-being. In order to improve public health outcomes, policymakers and practitioners should contemplate investing in and supporting initiatives of this nature. Although the results are encouraging, it is advisable to conduct additional research with a broader and more diverse sample, as well as longitudinal studies, in order to evaluate the long-term effects of these interventions and to investigate their efficacy in various contexts. In general, the “Chaitainya Utsav” event is a valuable model for future community-academy partnerships that are designed to promote community health and prevent substance abuse.

## Data Availability

All data produced in the present study are available upon reasonable request to the authors

